# Local habitual movement as a mechanism for *Schistosoma mansoni* transmission resurgence – a causal analysis

**DOI:** 10.64898/2026.04.06.26350236

**Authors:** Rivka M. Lim, Moses Arinaitwe, Simon A. Babayan, Andrina Nankasi, Alon Atuhaire, Annet Namukuta, Mwima Nicholas, Amy B. Pedersen, Joanne P. Webster, Poppy H.L. Lamberton, Jessica Clark

## Abstract

**Background/aims:** The World Health Organization (WHO) aims to eliminate schistosomiasis as a public health problem (EPHP) across 78 endemic countries by 2030. However, for low-prevalence settings that reach EPHP, guidance on managing transmission to maintain EPHP or move towards Interruption of Transmission (IoT) is limited, partly due to insufficient evidence on drivers of resurgence. In Uganda, some communities inland from Lake Victoria have achieved EPHP for *Schistosoma mansoni* but not progressed to IoT. This study explored whether routine, short-range travel to the highly endemic lake could sustain transmission in these settings.

**Methods:** We conducted a cross-sectional study in five Ugandan villages ∼5 km from Lake Victoria. Parasitological data were collected using Kato-Katz and Point-of-Care Circulating Cathodic Antigen tests, alongside questionnaires on lake travel from 585 individuals aged 1–91 years. A structural causal model estimated the total and direct effects of travel frequency, activity type, water contact duration, and drug treatment history on infection. Bayesian regression models and counterfactual simulations predicted infection under hypothetical interventions.

**Results:** Reaching IoT in low-risk villages may be undermined by habitual, short-range travel to high-risk sites, driven by the nature and duration of lake contact. Daily lake travel caused a 1.7-fold increase in odds of infection, while occupational activities caused a 3.4-fold increase compared with no lake activity. Counterfactual analysis showed that removing lake contact duration most reduced infection risk among moderate-frequency travellers, while daily travellers showed smaller changes, and some transmission persisted among individuals with little or no lake contact. Simulations demonstrated that modifying lake contact behaviours could reduce individual infection risk but had limited population-level impact.

**Conclusion:** These findings indicate that targeting only high-risk villages or individual behaviours is unlikely to achieve sustained, wide-spread IoT, underscoring the need for integrated control strategies that account for mobility, behaviour, and local transmission ecology.

**Key questions:** *What is already known on this topic?:*

- There is considerable guidance provided by the WHO for achieving EPHP, based on a wealth of scientific evidence. However, evidence to support the development of resources for areas that have reached EPHP is limited, particularly with regards to when and how to scale back mass drug administration (MDA), whilst mitigating the risk of resurgence.
- Simulation studies suggest that reducing treatment in low-prevalence settings may trigger a return to higher transmission, but the mechanisms sustaining residual infections in these contexts remain unclear and are often identified through statistical associations rather than through robust causal analyses. As such, there is a lack of evidence identifying drivers of resurgence.
- Human mobility is recognised as a potential driver of schistosomiasis transmission. However, previous studies have focused on long-distance migration or spatial risk modelling to predict future impacts of movement. The impact of individual-level frequent, habitual, local travel between low- and high-prevalence areas on transmission and resurgence has received comparatively less attention.

*What this study adds?:*

- This study provides causal evidence, rather than just statistical associations, that daily travel from low-prevalence inland villages to high-prevalence lakeshore landing sites increases the probability of *S. mansoni* infection risk and intensity, largely through activity type and duration of water contact.
- Counterfactual and mechanistic simulations show that reducing such travel would lower infection risk, but transmission persists among individuals with little or no lake contact, suggesting that without MDA, travellers may sustain infection in their home villages.

*How might this study affect research, practice or policy?:*

- Our findings suggest that low-prevalence communities should not automatically receive less frequent MDA.
- Equally, nor should communities who have achieved EPHP see their MDA frequency reduced without understanding sources of and plans for mitigating re-introduction of infection.
- Surveillance in such settings should attempt to capture mobile individuals and use more sensitive diagnostics to avoid underestimating residual risk.
- That communities are not isolated in space should be recognised when developing policy guidance, due to the risk of infection resurgence.
- Research and policy should continue to leverage causal approaches to clarify the mechanisms sustaining transmission in low-risk communities.

## Introduction

Schistosomiasis is a parasitic disease caused by trematode worms of the genus *Schistosoma*. In sub-Saharan African, where over 90% of human cases occur, the dominant species are *S. mansoni* and *S. haematobium* (causing intestinal and urogenital disease, respectively). Transmission occurs through contact with freshwater that has been contaminated through open defecation or urination, or inadequate containment of urine or stool infected with parasite eggs. These eggs then hatch and infect intermediate-host freshwater snails, where they develop into larvae which are infective to humans when released from the snails. Schistosomiasis transmission is highly localised, with infection prevalence and intensity varying substantially even between neighbouring communities (1). In response to the significant health burden posed by schistosomiasis, the World Health Organization (WHO) recommends a coordinated set of interventions, including primarily mass drug administration (MDA) with praziquantel, in combination with health education, improved access to safe water and sanitation, and snail control (2,3). While these strategies have reduced prevalence in many settings, it remains unclear whether, when and how best to scale back MDA, with the 2021-2030 Neglected Tropical Disease Roadmap, developed by the WHO, highlighting the need to understand how to monitor for, or what mechanisms could drive resurgence (4).

The WHO outlines three sequential programmatic goals to guide control efforts against schistosomiasis: 1) morbidity control; 2) elimination as a public health problem (EPHP); and 3) interruption of transmission (IoT) (2). An area is defined as having achieved these goals by using infection intensity, which in the case of intestinal schistosomiasis is quantified by egg counts generated using the Kato-Katz technique (5). For intestinal infection, a community is considered to have achieved morbidity control when <5% of school aged children (SAC) are heavily infected (defined as ≥400 eggs per gram of faeces (EPG)), EPHP is considered to have been achieved when <1% of SAC are heavily infected, and IoT is defined as having no autochthonous human or snail cases for five consecutive years (2). Progress towards these targets is assessed through combined surveillance, monitoring egg-based infection intensity and prevalence (5), and in some cases supported by Point-of-Care Circulating Cathodic Antigen tests (POC-CCAs) (6).

Whilst the WHO provides clear guidance for disease management and achieving EPHP in moderate and high prevalence settings (2), there are limited resources EPHP has been achieved. However, simulation models have indicated that when prevalence is <1% (by Kato Katz), residual persistent transmission can lead to resurgence without sufficient treatment (7) and surveillance (8). Furthermore, the impact of this is evidenced by resurgence in China following the reduction of interventions (9) and in Zanzibar after a one-year pause in MDA (10). Despite this risk, there are currently no robust post-EPHP surveillance and monitoring protocols (11), nor defined indicators for when to stop MDA or surveillance. Without clear guidance on how to detect early signs of resurgence or respond effectively, areas approaching EPHP or IoT, risk losing any progress made. To mitigate this risk, it is essential to identify the mechanisms that sustain low-level transmission and may drive resurgence, to inform surveillance and treatment strategies critical to long-term elimination and interruption success.

Communities living on the shores of Lake Victoria, Uganda tend to rely heavily on the lake for daily life. However, despite two decades of MDA, many continue to experience high rates of *S. mansoni* transmission (12,13). In contrast, some inland communities have shown reductions in prevalence, with some reaching EPHP, albeit without achieving IoT (14). This raises the question of whether inland communities which have reached EPHP, fail to progress to IoT due to their proximity to high-endemicity regions, and specifically whether transmission is sustained through habitual human movement between these low- and high-risk locations.

Local habitual human mobility between low and high transmission regions has received comparably less attention than long-term/distance human migration or movement (15–18). Where previous studies have identified human mobility as a key factor shaping the spatial dynamics of schistosomiasis transmission, they have largely relied on indirect evidence, such as spatial models of risk, or ecological correlations, to infer associations between movement and infection (19–21) and have not focussed on the impact of habitual movement between areas which have achieved EPHP and those where endemicity remains high. Existing control strategies often operate by administrative units, assuming that populations are geographically isolated, and as such likely obscuring the possibility that habitual local mobility could mean individuals act as a conduit for sustained transmission. This raises another important question of whether certain people should, or can, be prioritised for surveillance or treatment during the elimination phase, as a function of their connectivity to high-risk locations, to prevent re-establishment of local transmission.

Additionally, although travel-related behaviours such as water contact and time spent in the lake are known risk factors for schistosomiasis transmission (22), there is little evidence on whether particular individuals, or their activities, are the cause of this risk. Understanding whether routine travel directly increases infection risk, or whether it acts indirectly through other behaviours is essential to inform targeted elimination strategies. Traditional epidemiological analyses often describe statistically significant associations between exposures and outcomes but cannot determine whether those exposures drive the outcomes (i.e. evidence causality). In contrast, causal inference methods, like structural causal modelling (SCM), are specifically used to estimate the effects of exposures while accounting for confounding, mediation (where effects operate through intermediate variables), and complex interdependencies. These approaches are increasingly applied in infectious disease epidemiology to generate policy-relevant insights, where programmatic decisions must be made using observational data (23,24). By explicitly defining and testing assumptions regarding the ways different drivers might interact, and the pathways through which transmission is shaped, SCMs offer a rigorous framework to evaluate drivers of infection and support elimination planning.

The aim of this study was to quantify the causal effect of habitual, local travel to and from highly endemic Lake Victoria areas on *S. mansoni* infection risk in regions that have met WHO targets for EPHP. To do this, we applied a Bayesian causal inference modelling framework to highly detailed behavioural and parasitological data collected from five communities, to estimate the direct and indirect effects of travel-related behaviours on infection risk. Understanding these patterns of movement and their relationship to infection can offer valuable insight into whether such individuals may be contributing to ongoing transmission that could be hindering progress toward IoT. Clarifying this relationship will support the development of efficient and effective surveillance strategies, improve disease resurgence detection, and ultimately contribute to achieving IoT goals.

## Methods

### Ethics

Informed written consent via signature or thumbprint was gathered prior to any data collection for each adult, or from a legal guardian of those under the age of 18. In addition to guardian consent, those aged eight to 17 gave informed signed or thumb printed assent. All participants understood that they could leave the study at any point without losing access to treatment. Enrolled participants were allocated a numerical ID which pseudo-anonymously linked parasitological and travel data.

### Public involvement

Community members were first engaged during a sensitisation visit led by Ministry of Health team. The research questions were developed from policy priorities to understand community experiences and practices related to schistosomiasis and its treatment, particularly in low-risk areas where data are limited. While the public did not directly inform study design, outcome measures, or recruitment, participation was voluntary following the sensitisation meetings, during which questions were addressed before consent. Feasibility was thus dependent on community acceptance. The pilot questionnaire was refined based on volunteer feedback.

### Data collection

Five villages each ∼5 km from Lake Victoria, Uganda were selected: B, Nam, Nan, Nw and S (Figure 1) (see supplementary data “Village selection” for full selection protocol). Individuals aged one-91 years old were recruited assisted by the village health team. The study took place in each village for one week, from October 2021-December 2021.

**Figure 1.**
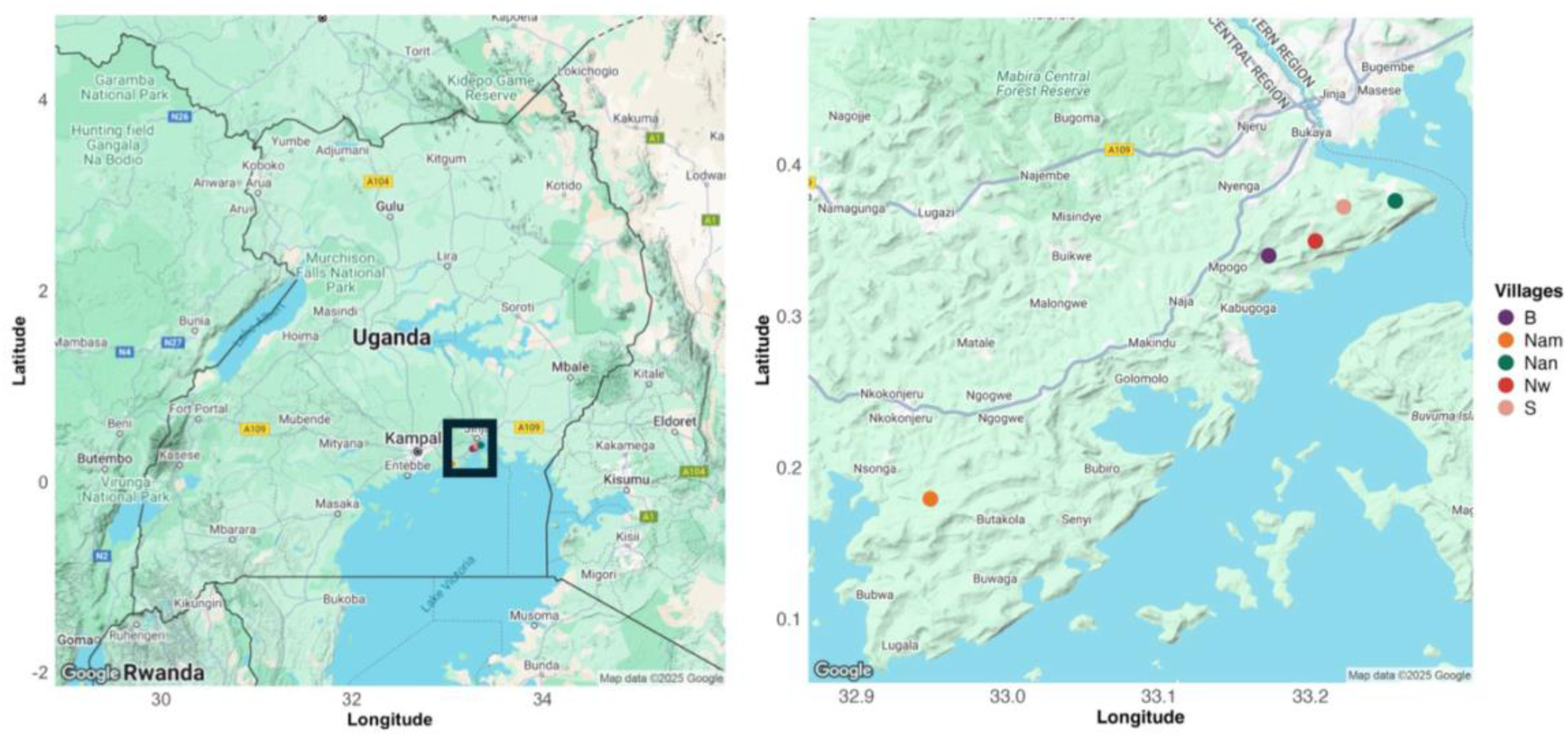
Map of the study area showing the locations of the five participating villages along the northern shore of Lake Victoria, Uganda. Each point represents one village, colour-coded by name: B (purple), Nw (red), Nam (orange), Nan A (green), and S (pink). The left panel provides a broader map of Uganda, with a black rectangle indicating the location of the zoomed-in map shown in the right panel, which displays a detailed view of the study area.

Three stool samples were collected per person over three days and duplicate Kato-Katz examined by experienced technicians, under a microscope, for *S. mansoni* eggs (5). One urine sample per person was tested using POC-CCAs, using the 1-10 G-Score system (25). Kato-Katz have 100% specificity but POC-CCAs were included to improve sensitivity in low prevalence settings (26). A participant was considered positive for *S. mansoni* if at least one egg was observed in a stool sample, or if negative by eggs but a POC-CCA ≥G3 (27,28). Prevalence was calculated by dividing the number of positive people by the full sample size. Individual mean infection intensity (EPG) was calculated as the mean slide count multiplied by 24. Population-level mean EPG was calculated removing individuals with EPG=0.

Human mobility and water contact were assessed through a questionnaire administered once during the study period. Questionnaires were conducted in the local language by trained fieldworkers and answers recorded on tablets with KoboToolBox. Respondents were asked their age, sex, and current village, as well as how many times they had travelled to Lake Victoria in the past three months: i) never, ii) just once, iii) once a month, iv) twice a month, v) once a week, vi) twice a week, or vii) daily; what activities they engaged in upon arrival and how long they were in the water. Age was categorised into three programmatically relevant groups: PSAC (0–5 years), SAC (6–15 years), and adults (>15 years). We explored an alternative four-level specification including young adults (15-25 years) in sensitivity analyses. As effect estimates for young adults were comparable to those for older adults, with no improvement in model fit or predictive accuracy, the three-level age variable was retained for the analysis. Questions regarding MDA participation were also included.

Participants reported a wide variety of activities undertaken at the lake. These activities were grouped into i) domestic, ii) occupational, iii) recreational, and iv) trade/visit and were used in all models. A complete list of activity responses and their groupings is provided in Table S1.

### Directed acyclic graph construction

All analyses and visualisations were executed using R (v. 4.4.3). A directed acyclic graph (DAG) was constructed to represent hypothesised causal relationships influencing the link between an individual’s infection status and travel to a highly endemic location. The DAG was then used to inform the structure of the final model used for analysis. This DAG included travel frequency to Lake Victoria, water contact duration at Lake Victoria, activity type at the lake, MDA history, and demographic covariates: age, sex, and village of residence (Figure 2A). The structure of the DAG i.e. direction of the edges and which variables interact, was informed by a combination of domain knowledge, published literature, and expert opinion of schistosomiasis transmission dynamics, local environmental conditions, and behavioural patterns (29). The DAG aimed to visually represent a biologically plausible and theoretically grounded causal framework. The DAG was visualised using the R package dagitty (30). The structure of the DAG was validated against the observed data by testing the implied conditional independencies between the variables (see supplementary data “Model validation and restructuring” for a full explanation).

**Figure 2.**
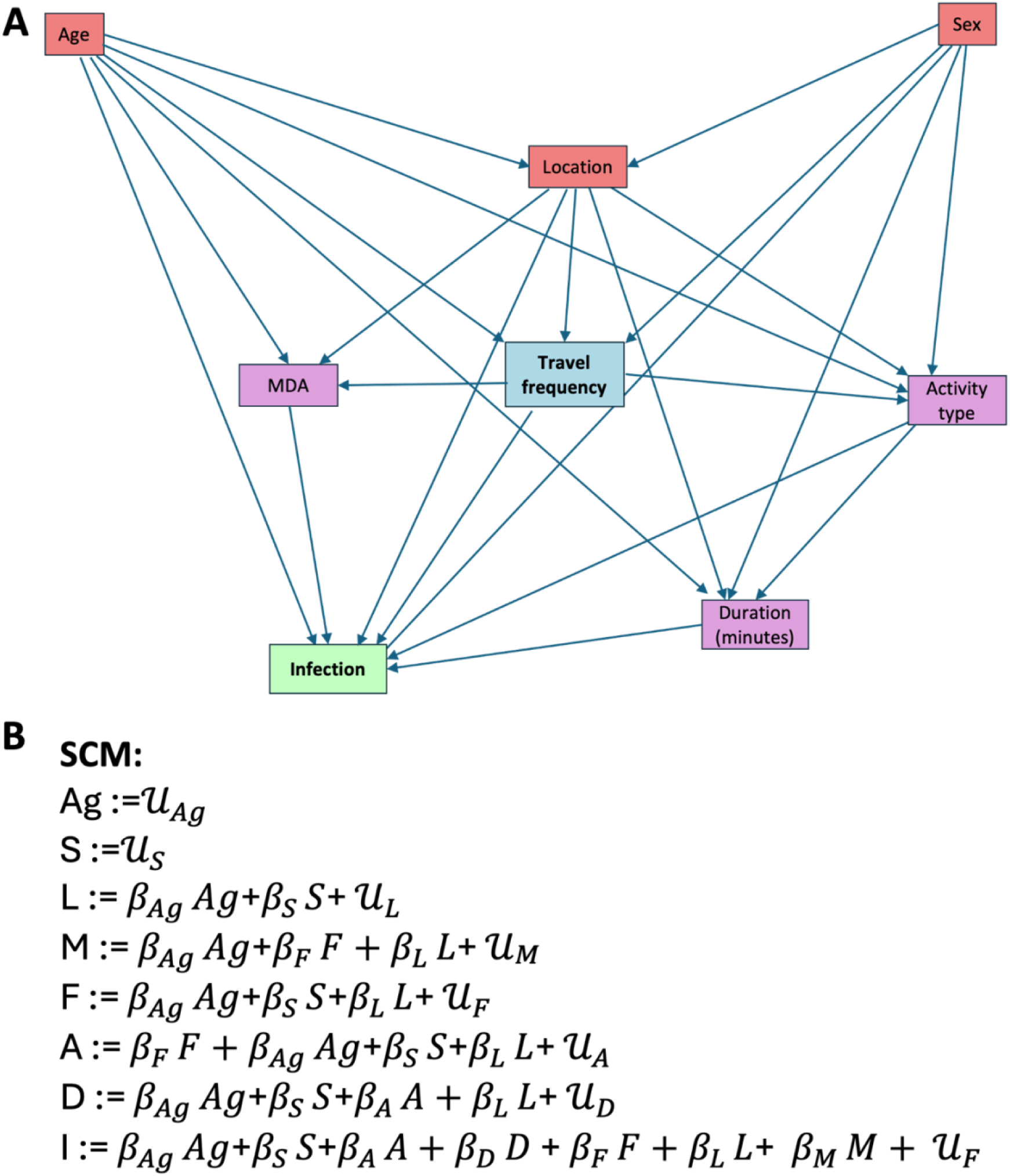
Directed acyclic graph (DAG) representing hypothesised causal relationships between variables and *Schistosoma mansoni* infection (A), with structural equations for each node (B). (A) The DAG outlines the assumed causal structure linking demographic, behavioural, and environmental variables to infection. Arrows represent direct causal effects. Key exposure variables include travel frequency, activity type, and duration of water contact, with potential confounders such as age class, sex, and location. (B) Structural equations define the dependencies of each variable, with terms representing regression coefficients (β), unobserved residual error (U), and observed covariates. Ag = age class, S = sex, L = location, M = MDA, F = frequency of travel, A = activity, D = duration in the lake, I = infection.

### Structural causal model

From the fully validated DAG, a SCM was constructed for analysis. Each variable was expressed as a function of its direct causes (parents in the DAG) and an independent noise term (Figure 2B). These functions define the joint distribution of all variables in the system, informing the underlying causal structure. Minimal adjustment sets for each exposure-outcome relationship were estimated from the SCM to ensure an unbiased estimation of total and direct effects (Table S2) (31,32). These adjustment sets were formulated using causal graph theory principles based on chains, forks and colliders (33). To account for uncertainty and provide probabilistic estimates of causal effects, we used Bayesian linear regression models. Each edge in the DAG was parameterised using these Bayesian models. Effects were considered supported if the 95% credible interval (95% crI) for the effect between variables did not include zero after adjusting for the conditioning set identified by the DAG structure. Using these appropriate adjustment sets we estimated total and direct effects of key causes of infection status and intensity, fitting models for each of the four primary causes: travel frequency, water contact duration, MDA history and activity type. Bayesian linear models were fitted, and posterior predictive checks conducted using the *brms* package in R (34). Binary infection status (*S. mansoni* presence or absence) was modelled using a Bernoulli distribution with a logit link. Egg burden (measured as mean EPG, among egg-positive individuals only) was modelled using a Gamma distribution with a log link, given its positive and right-skewed distribution. After carrying out sensitivity analyses for choice of coefficient priors (supplementary data “Sensitivity analysis of priors for the coefficients” and Figure S1), weakly informative priors were chosen and differed by model family. Priors for each of the models are shown in supplementary data (Table S3). Posterior distributions were drawn using four chains of 4000 to 10,000 iterations each (typically with 30% warm-up), depending on model complexity.

### Counterfactual interventions

To simulate counterfactual interventions, we used posterior draws from the total effect models to generate predicted probabilities of infection and predicted mean EPG under both observed and intervened exposure conditions. For each intervention, we selected a target exposure variable (travel frequency, water contact duration and activity type) and modified it to reflect the desired intervention. Where an exposure had downstream effects on other variables in the DAG (Figure 2A), these variables were also modified, for example if someone doesn’t travel to the lake, duration in the lake would also be set to zero.

The original model, fitted with the unmodified factual dataset using the appropriate adjustment sets (Table S2) was then applied to both the factual and counterfactual datasets to obtain posterior predictions. This preserved the natural variation in covariates needed to estimate *P*(*Y*|*X*) when applying the do-calculus, where *Y* denotes the outcome (binary infection status or mean EPG) and *X* the exposure of interest.

This approach enabled us to estimate how infection risk would change under hypothetical interventions while avoiding post-treatment bias. We applied this approach to different scenarios: A) **Travel frequency:** setting all daily travellers to no travel, examining (i) change in infection probability for the entire population, and (ii) change only for daily travellers; B) **Activity type:** (i) changing all “occupation: activity to “none” and examining infection probability for the whole population and occupational travellers, (ii) changing all “domestic” activity to “none” and examining mean EPG for the whole population and for domestic-activity individuals only; and C) **Water contact duration:** setting duration to zero, examining (i) change in infection probability for the entire population, and (ii) change only for those who had reported lake contact.

For each scenario, posterior predictive distributions were summarised as means across individuals for each posterior draw and visualised under observed and counterfactual conditions using histograms. Presenting full distributions rather than single point estimates allowed assessment of uncertainty, skew, and variability in predicted effects.

### Mechanistic counterfactual analysis

To examine whether the effect of removing lake contact duration varied by travel frequency, we conducted a mechanistic counterfactual analysis. Posterior infection risks were predicted under the observed data and under a counterfactual scenario where duration was set to zero for individuals who had reported >0mins in the Lake. Conditional average treatment effects were then estimated by computing mean infection risks within each travel-frequency group and taking the difference between counterfactual and observed values. This approach quantified how the causal effect of removing lake contact duration differed across levels of travel frequency, providing a sensitivity analysis of heterogeneity in the duration-infection pathway.

## Results

### Prevalence of *Schistosoma mansoni* infections

Community-wide prevalence of infection was 51% in Nan, 45% in B, 33% Nam, 30% in Nw and 27% in village S (mean=36%). No participant had an infection intensity of ≥400 EPG (Table S4). As the WHO defines EPHP as <1% of SAC with ≥400 EPG, all villages had achieved EPHP (2). For more detailed village, age, and sex level variation stratified by travel habits, see supplementary Data (Table S4 and Figure S2).

### Effect of travel frequency on infection

Among all participants, 56% (329/585) reported travelling to the lake at least once in the last three months (Figure S3). Of these travellers, 41% (134/329) were infected and 30% of those that did not travel to the lake were infected (105/256) (Table S5). Daily travel to the lake was reported by 24% of participants (138/585). The mean EPG in each group (travel and no travel) was 11.3 (sd = 22.2) and 11.6 (sd = 13.4) respectively.

The total effect of travel frequency on infection was estimated using the minimal adjustment set of confounders only (Table S2). There was a positive total causal effect for those who travelled once a month, twice a month, twice a week and daily to Lake Victoria in the last three months, with daily visits having a significant 1.7x effect on increasing infection risk (log odds= 0.55, 95% crI=0.06 to 1.05) (Figure 3A). When estimating the direct effect, there was no longer a significant effect of daily visits (Figure S4), with the activities classed as occupation now having the only significant effect in the model (Figure 2A). Using a separate model of EPG, the amount of travel undertaken did not significantly affect the EPG in either the total or direct models (Figure 3B and Figure S5).

**Figure 3.**
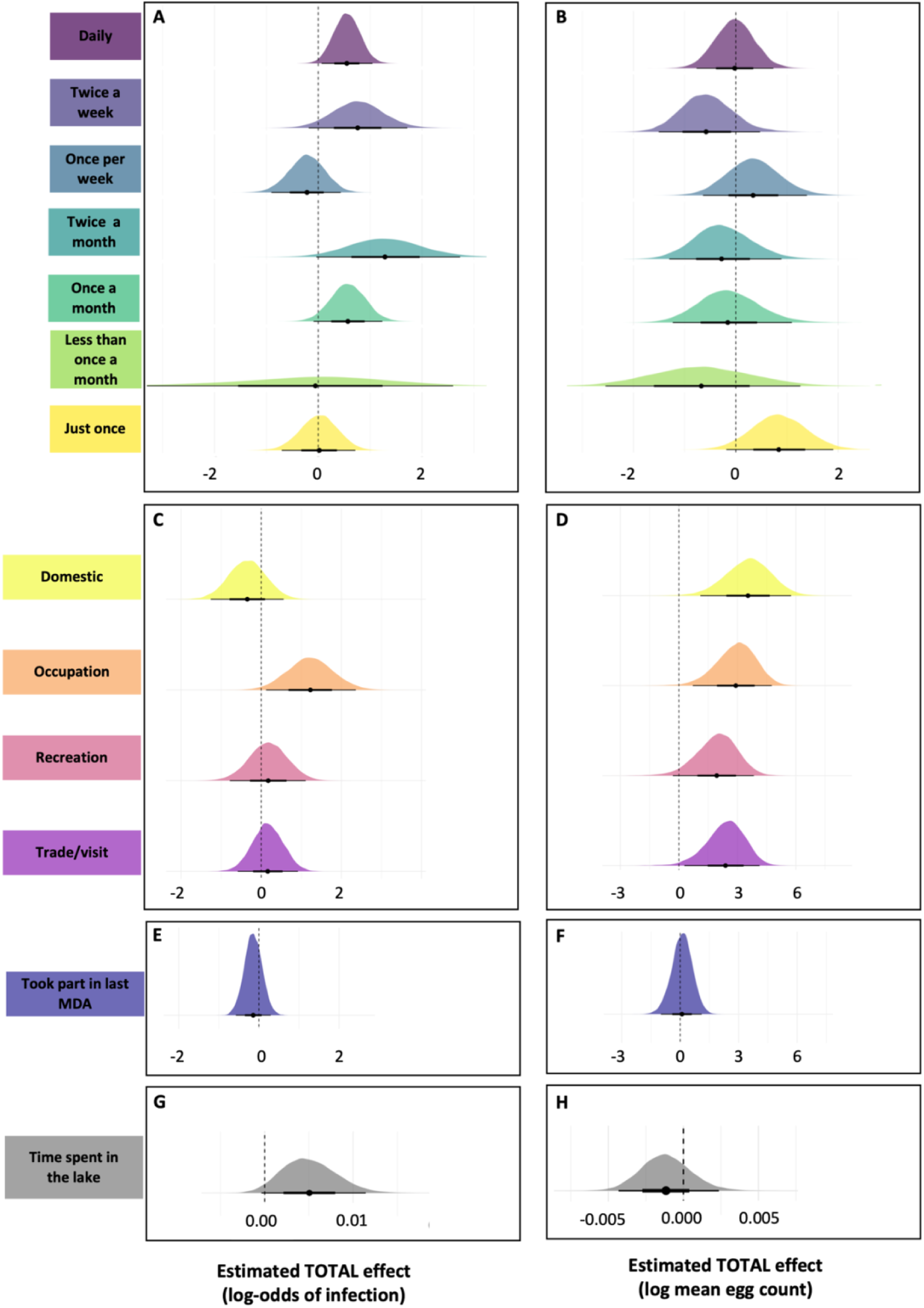
Estimated total effects of lake-related behaviours on *Schistosoma mansoni* infection and infection burden. Each pair of panels (left = infection probability; right = infection burden) shows posterior distributions from Bayesian regression models estimating the total effects of specific lake-related variables. <u>Panels A & B:</u> Self-reported travel frequency to Lake Victoria (relative to no travel) on (A) the probability of *S. mansoni* infection and (B) infection burden measured as eggs per gram (EPG) of stool). <u>Panels C & D:</u> Self-reported lake-related activity type on (C) probability of infection and (D) infection burden. <u>Panels E & F:</u> Participation in the most recent mass drug administration (MDA) on (E) infection probability and (F) infection burden. <u>Panels G & H:</u> Time spent in contact with lake water on (G) infection probability and (H) infection burden. Densities represent marginal posterior distributions of log-odds (infection) or log-mean EPG (burden). Thick and thin horizontal lines represent 66% and 95% credible intervals, respectively. The vertical dashed line indicates the null effect (log-odds or log-mean = 0). In panel A, travel frequency categories are ordered from highest (daily visits) to lowest (single visit). Observed infection prevalence and intensity by behavioural and exposure variables can be visualised on Figure S3.

### Effect of activity carried out whilst visiting Lake Victoria

When estimating the total effect of activity on infection, individuals reporting occupational water contact had higher log-odds of being infected compared to those with no lake contact (log-odds = 1.23; 95% crI= 0.12 to 2.36). Other activities were not significant predictors of infection risk (Figure 3C).

Including water contact duration in the model to estimate the direct effect reduced the estimated effect of occupational activity on infection (log odds = 0.91; 95% crI= –0.26 to 2.10), compared to the model without duration (Figure S6). Duration itself was positively associated with infection risk (log-odds = 0.01, crI=0.0 to 0.01), confirming partial mediation as outlined in the DAG (Figure 2A).

All activities were associated with a higher mean EPG compared to those reporting no lake-related activity. Domestic activities had the greatest effect with a 35x higher mean EPG than those reporting no activity at the lake (log odds= 3.55, 95% crI= 1.10 to 5.76) (Figure 3D). Adjusting for duration of lake exposure in a direct effect model, did not meaningfully alter the association between activity type and EPG in comparison to the total effect model (Figure S7), and duration was not a significant predictor.

### Effect of taking part in the last MDA

Across all villages, 73% (425/585) of participants said they took part in the last MDA. Details of responses to why a participant did not take part in the last MDA and what age groups did take part can be found in supplementary data (Tables S6 And S7). When modelling the effect of MDA participation on infection, the total and direct effects used the same adjustment sets, as there were no recorded mediators (Table S2). Individuals who reported participating in MDA had lower though non-significant log-odds of infection than those who did not (log-odds= –0.14; 95% crI = −0.57 to 0.29) (Figure 3E). There was no association between MDA and mean EPG (log-mean = 0.09; 95% crI= –0.99 to 1.11) (Figure 3F).

### Effect of time spent in Lake Victoria

The longest lake visits were for occupational activities (mean time spent (minutes) = 92, sd = 161), recreation followed (mean= 33, sd = 32), then trade/visiting (mean= 18, sd = 34), with domestic activities shortest (mean= 14, sd = 32). For daily visits, occupations were rare (n = 9) but longest (mean= 64, sd = 70), while domestic visits were most common (n = 73) and shortest (mean= 5, sd = 12) (Table S8).

When modelling the effect of time spent in Lake Victoria on infection, the total and direct effects used the same adjustment sets as there were no mediators (Table S2). The log-odds of infection increased by 1% (log-odds = 0.01, 95% crI= −0.0 to 0.01) for every additional minute (Figure 3G) (note that the lower bound is essentially on 0 suggesting an incalculably small possibility of 0, and so technically only a weekly positive association). While this effect is small per minute, it accumulates across longer durations, for example, an additional 60 minutes corresponds to approximately 82% higher odds of infection than someone who spends no time in the lake (assuming linearity) (log-odds of (0.01* 0.6)). There was no effect of duration on mean EPG (log-odd = −0.0, 95% crI= −0.0 to 0.0) (Figure 3H).

### Counterfactual interventions

The first counterfactual simulation removed daily lake travel, reducing infection probability by 3% in the whole population from 36% (95% crI: 32–40%) to 33% (95% crI: 29–38%) (Figure 4A) and by 12% among daily travellers from 41% (95% crI: 33–50%) to 29% (95% crI: 23–36%) among daily travellers (Figure 4B).

**Figure 4.**
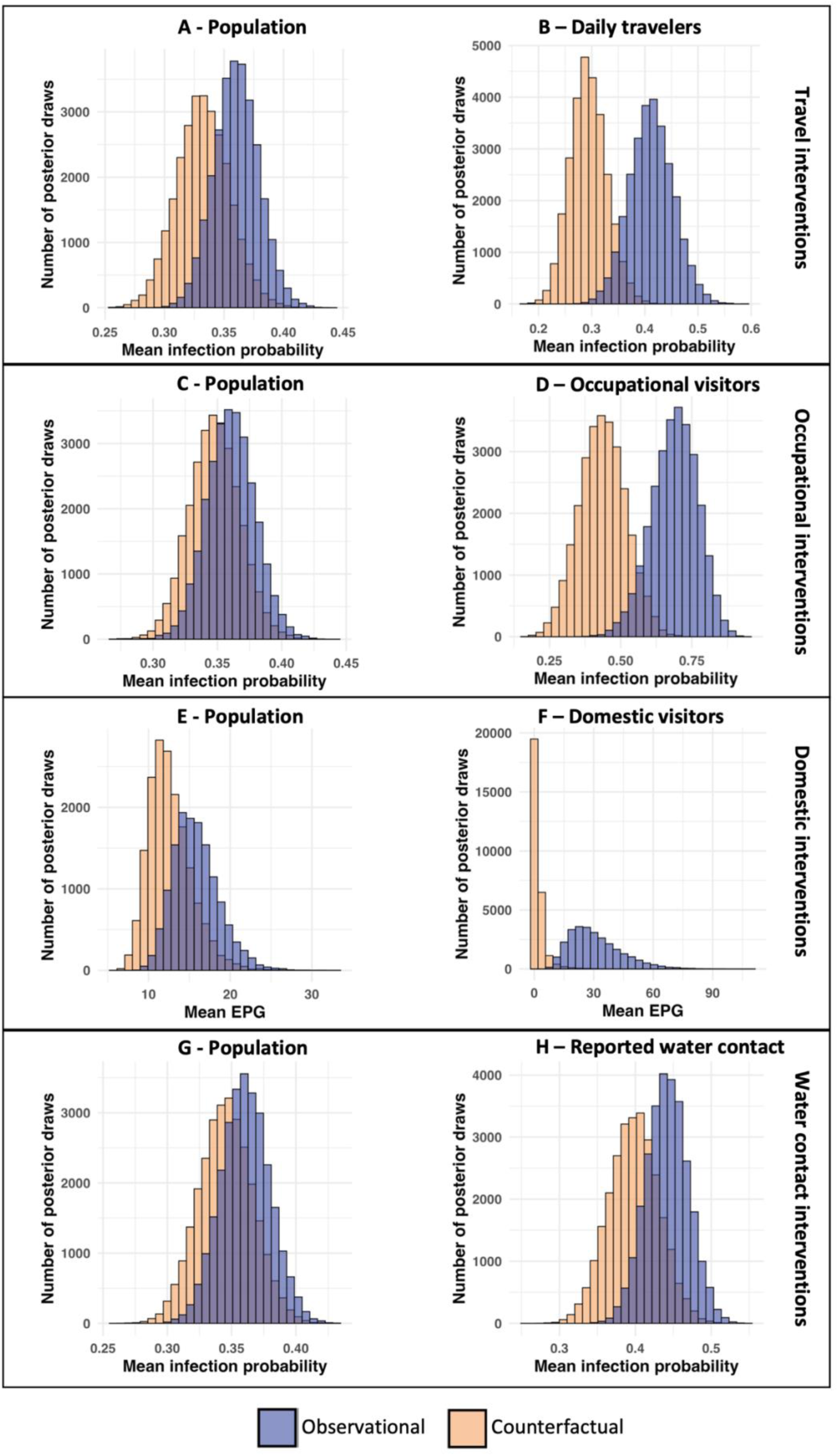
Counterfactual predictions of *Schistosoma mansoni* infection probability under reduced travel and water contact scenarios. Posterior distributions of predicted infection probabilities for selected subgroups under observed (blue) and counterfactual (orange) conditions, based on a Bayesian logistic regression models. **A and B)** Travel interventions: the predicted infection probability of no daily travel on the entire population (A); and only daily travellers (B). **C and D)** Occupational interventions: the predicted infection probability of no one doing occupational activities on the entire population (C); and only those going for occupational tasks (D). **E and F)** Domestic interventions: the predicted mean eggs per gram of no one doing domestic activities on the entire population (E); and only those going for domestic tasks (D). **G and H)** Water contact interventions: the predicted infection probability of no water contact on the entire population (G); and only those who reported making contact (H). Distributions represent posterior samples of individual infection probabilities aggregated across the sample population.

Removing occupational lake visits reduced infection probability by 1% in the whole population from 36% (95% CrI: 32–40%) to 35% (95% crI: 31–39%) (Figure 4C) by 26% among those reporting occupational visits, from 70% (95% crI: 52–83%) to 44% (95% crI: 28–59%).

Eliminating domestic water activities reduced mean infection intensity from 16 EPG (95% crI: 11–22) to 12 EPG (95% crI: 9–19) in the overall population (Figure 4E) and from 29 EPG (95% crI: 12–62) to 1 EPG (95% crI: 0.2–11) among those reporting domestic activities (Figure 4F).

Setting lake contact duration to zero reduced infection probability from 36% (95% crI: 32–40%) to 35% (95% crI: 31–39%) in the overall population (Figure 4G) and from 44% (95% crI: 39–50%) to 40% (95% crI: 33–46%) among those reporting contact (Figure 4H), a reduction of 1% and 4%, respectively.

### Mechanistic counterfactual: contact duration by travel frequency

To examine whether the effect of removing lake contact duration varied across levels of travel frequency, we estimated conditional average treatment effects (CATEs). Reductions in infection probability were minimal among non-travellers and infrequent visitors (≤ once a month), highest amongst those travelling twice per month to twice per week (difference in infection risk of six-10 percentage points – Table 1). Daily travellers, while most at risk overall (Figure 3), showed comparatively small reductions (three percentage point reduction – Table 1).

**Table 1.** Contact-only estimated change in infection probability under do(Duration = 0)

| Travel frequency | Median $\Delta$ (risk difference) | 95% CrI (lower, upper) |
| --- | --- | --- |
| Non-travellers | -0.030 (≈ 3%) | -0.071, 0.002 |
| Just Once | -0.029 (≈ 3%) | -0.065, 0.002 |
| < Once per month | -0.034 ($\approx 3\%$) | -0.079, 0.003 |
| Once per month | -0.038 ($\approx 4\%$) | -0.088, 0.003 |
| Twice per month | -0.102 ($\approx 10\%$) | -0.213, 0.011 |
| Once per week | -0.058 ($\approx 6\%$) | -0.133, 0.004 |
| Twice per week | -0.082 ($\approx 8\%$) | -0.158, 0.008 |
| Daily | -0.031 ($\approx 3\%$) | -0.071, 0.002 |

## Discussion

This study aimed to test whether habitual movement of people living in low *S. mansoni* endemicity areas to high-transmission sites around Lake Victoria, Uganda, could be sustaining transmission in surrounding inland communities that have reached the WHO-defined threshold for EPHP (2). Our analysis revealed that frequent travel, particularly daily travel is a plausible and quantifiable pathway for sustaining *S. mansoni* transmission in low-prevalence settings. Critically, our results indicate that multiple overlapping exposures contribute to infection. It is therefore unlikely that transmission can be controlled by a single intervention, highlighting the need for a systems-thinking approach that recognises behavioural, environmental, and structural factors influencing transmission dynamics in EPHP settings.

To explore how behavioural interventions might influence infection risk and assess potential resurgence pathways, we simulated a series of counterfactual scenarios using the fully resolved SCM. The aim was not to advocate for behavioural restrictions, but to identify groups who might benefit most from enhanced surveillance or targeted treatment. Across all scenarios, predicted infection probabilities and mean EPG were lower than in the observed data. The largest reductions in infection were consistently observed within the specific subgroups targeted by each intervention, with relatively small effects at the population level. At the individual scale, such interventions could be considered successful, reducing infection probability by 12% in daily travellers, 26% in occupational visitors, and lowering mean EPG in those undertaking domestic water activities from 29 to 1. These changes would not only improve individual health outcomes but also reduce the likelihood of those individuals contributing to contamination of home-village water sources in the future. However, the insubstantial impact at the population level highlights a limitation of targeting individual risk factors to control transmission in established transmission settings.

Building on these modelled insights, the empirical data show that daily travel to Lake Victoria was associated with a 1.7-fold higher probability of infection compared with no travel, confirming a causal link between travel frequency and infection. When potential mediators were included, this association became non-significant, indicating that the elevated risk among daily travellers is largely explained by lake-related activities and time spent in the water. Participants involved in occupational water contact had a 3.4-fold higher infection probability than those reporting no lake activity, spending on average 92 minutes in the lake compared with 14 minutes for domestic activities. Although domestic activities were not linked to higher infection probability, they were strongly associated with higher mean EPG among those infected, possibly reflecting occasional high-risk behaviours such as washing clothes (22% of domestic cases) in shallow shoreline waters where Biomphalaria snails and cercarial densities are highest. Weekly travellers showed no elevated infection risk despite moderate exposure durations (∼39–40 minutes), likely due to the low proportion engaged in occupational contact (4%).

Mechanistic counterfactual analysis, simulating zero minutes of lake contact while stratifying by travel frequency to assess the direct contribution of contact duration independent of mobility behaviour, revealed the greatest reductions in infection probability among those travelling twice per month, whereas daily travellers, the group at highest overall risk showed smaller changes, likely reflecting residual infection in this group. Taken together, these findings indicate that infection risk is shaped by the frequency, nature, and duration of water contact, and that some transmission persists among individuals with little or no lake contact. This further supports the earlier evidence that multiple overlapping exposures contribute to infection risk. The detection of infections among participants who never travelled to the lake supports the existence of inland transmission foci sustained by parasite importation from mobile individuals. Frequent travellers therefore appear to play a pivotal role in seeding infections within their home communities through routine water contamination, allowing transmission to persist among non-travellers and across the wider area. This connectivity blurs the boundaries between “high-risk” and “low-risk” villages. Although all five study villages met EPHP criteria, with no heavy-intensity infections and low mean egg counts among egg-positive individuals (11 EPG, SD = 20), these low-intensity but persistent infections have important public health implications, highlighting the risk of underestimating transmission potential and resurgence in post-EPHP settings, suggesting that reliance on egg counts alone underestimates the true reservoir.

We therefore suggest that control in settings which have achieved EPHP require a systems-based approach that addresses more than just the proximal drivers of transmission (e.g. human behaviour and environmental reservoirs) to acknowledge the multiplicative effects of distal drivers that are often socio-political and structural in nature (35). Our statistical framework provides a tool to assess the potential impact of such strategies without relying on large, highly parameterised dynamic models, which require extensive, context-specific data. By linking empirical behavioural data to infection outcomes, this approach can help understand mechanistic pathways which could lead to resurgence. Looking forward, this framework could also inform adaptive post-EPHP surveillance and intervention strategies, enabling programmes to anticipate and respond dynamically to early signals of re-emergence.

### Limitations

This study relied on self-reported behaviour from questionnaires, rather than direct observations. While direct observation would offer more accurate behavioural data, it was not feasible at the scale of this study. Misreporting is a known issue with self-reports (36), and one such example emerged in our data when participants were asked about water contact at or near their homes. Technicians working on this project had noted that many individuals used nearby swampland as a local water source, and so a set of questions was included to distinguish infection risk from local versus Lake Victoria exposure. However, upon examination of the reported home water sources, 50% were found to be on the shores of Lake Victoria, suggesting the question was misunderstood. As a result, these data, and responses to all home water contact questions, were excluded from the analysis due to concerns over validity.

Another limitation relates to interpretation of MDA participation. Although 74% of participants reported taking part in the most recent MDA, the effect of treatment on infection risk was weak, and no effect was observed on infection burden. This could be due to rapid reinfection, or low drug efficacy, but may also reflect overreporting of participation. This is a well-documented issue in qualitative research, where answers align with what the participant believes is most desirable to the researcher (37). Furthermore, 50% of PSAC reported taking part in last MDA, although were not eligible due to a lack of paediatric praziquantel at the time. Attempts were made to retrieve the district reports on MDA, but it was not possible, therefore the validity of self-reporting is without direct confirmation (e.g. drug registers).

We hypothesised that high levels of habitual travel could also increase the risk of missing MDA rounds, and our data supported this, with 28 individuals reporting they missed the last MDA due to travel. However, if missing MDA is possible, so is missing surveys such as our own study, and such individuals may be systematically under-represented in surveillance and/or research activities, and may bias both epidemiological estimates and intervention evaluations (38). In our setting, this implies that the infection risk and MDA non-participation associated with high travel frequency may be underestimated, precisely because the most mobile individuals may have been absent during data collection.

### Conclusions

The findings presented here suggest that, in regions which have achieved EPHP, a small subset of individuals engaging in high-risk behaviours may help to sustain local prevalence, particularly occupational lake contact and habitual travel, and may serve as a bridge in maintaining infection within otherwise low-risk groups. Counterfactual simulations showed that while targeted interventions substantially reduce risk in individuals, their effect at the population level is negligible, indicating that such approaches alone are insufficient. Preventing resurgence will therefore require a systems-based strategy that integrates human, environmental, structural, and ecological drivers of transmission to safeguard long-term gains and achieve durable interruption of *S. mansoni* transmission.

## Data Availability

All data produced are available online at https://github.com/iamjessclark/travelStudySchisto

## Data Availability

All data produced are available online at https://github.com/iamjessclark/travelStudySchisto

## Acknowledgements

We would like to thank the communities of, and individuals in, the study villages for participating in this study.

## Data Availability

https://github.com/iamjessclark/travelStudySchisto

## Ethical Approval Statement

Study protocols were strictly adhered to following approval by the University of Glasgow MVLS ethics committee (200160068), Ministry of Health Uganda, Vector Control Division research ethics committee (VCDREC/062) and Uganda National Council for Science & Technology (UNCST-HS 2193).

## Supplementary Figures and Tables

**Table S1.** Participants responses to what activity they did at Lake Victoria put into groups for statistical analysis

**Table S2.** Minimal adjustment sets used in total and direct effect models by exposure.

**Figure S1.** Total effects of activity on *Schistosoma mansoni* infection burden.

Posterior estimated total effects of lake-related activities, on the log mean of *S. mansoni* eggs per gram of stool. The model included fixed effects for activity type (domestic, occupational, recreational, trade/visit), and was adjusted for age class, sex, and location and duration.

Posterior densities represent marginal distributions of estimated effects, with the vertical dashed line indicating the null (log = 0).

Moderately informative coefficient priors of Normal(0,5)

Conservative coefficient priors of Normal(0,2)

**Table S3.** Model parameters and prior distributions.

**Table S4.** *Schistosoma mansoni* prevalence and mean eggs per gram (EPG) by travel frequency to Lake Victoria, Uganda

**Table S5.** Frequency of Lake Victoria travel and distribution of reported activities, stratified by age group, sex, and village.

**Figure S2.** Prevalence of *Schistosoma manson*i by village and Sex. Prevalence measured by a positive Kato Katz from three duplicate slides or if a POC-CCA test had a G-score ≥3. F = female and M = male.

**Figure S3.** Observed infection prevalence and intensity by behavioural and exposure variables.

Observed infection prevalence (top and middle rows) and infection intensity (bottom row) plotted against behavioural and exposure variables included in the structural causal model. Bars and ribbons show observed mean prevalence with 95% binomial confidence intervals. Points represent individual infections (1 = infected, 0 = uninfected), shown with jitter for visibility.

**(A)** Infection prevalence by travel frequency to Lake Victoria (visits in the past three months).

**(B)** Infection prevalence by participation in the most recent mass drug administration (MDA).

**(C)** Infection prevalence by activity type at Lake Victoria.

**(D)** Infection prevalence by time spent in the lake (binned durations).

**(E)** Infection intensity (EPG, log₁₊ scale) by time spent in the lake (continuous).

**(F–H)** Infection intensity (EPG, log₁₊ scale) by travel frequency (F), MDA participation (G), and activity type (H).

**Figure S4.** Direct effect of travel frequency on log odds of infection with *Schistosoma mansoni*.

Posterior distributions for the estimated direct effects of self-reported travel frequency to Lake Victoria on the probability of infection, relative to individuals reporting no travel in the preceding three months (reference category). Estimates are log-odds coefficients from a Bayesian logistic regression model with a Bernoulli likelihood and logit link function. The model included fixed effects for travel frequency, age class, sex (binary), and location and the mediator’s activity and MDA. Densities represent marginal posterior distributions for each travel frequency level, with the vertical dashed line indicating the null effect (log-odds = 0). Travel frequency levels are ordered from highest (92 days) to lowest (1 day) reported visits to the lake.

**Figure S5.** Direct effect of travel frequency on log mean on burden of *Schistosoma mansoni* eggs per gram in stool.

Posterior distributions for the estimated direct effects of self-reported travel frequency to Lake Victoria on mean eggs per gram of stool, relative to individuals reporting no travel in the preceding three months (reference category). Estimates are log mean coefficients from a Bayesian logistic regression model with a gamma likelihood and log link function. Densities represent marginal posterior distributions for each travel frequency level, with the vertical dashed line indicating the null effect (log-odds = 0). Travel frequency levels are ordered from highest (92 days) to lowest (1 day) reported visits to the lake.

**Figure S6.** Direct effects of activity on *Schistosoma mansoni* infection probability.

Posterior distributions for the estimated direct effects of lake-related activities, the log odds of infection. The model included fixed effects for activity type (domestic, occupational, recreational, trade/visit), and was adjusted for age class, sex, and location and duration. Posterior densities represent marginal distributions of estimated effects, with the vertical dashed line indicating the null (log = 0).

**Figure S7.** Direct effects of activity on *Schistosoma mansoni* infection burden.

Posterior distributions for the estimated direct effects of lake-related activities, on the log mean of *S. mansoni* eggs per gram of stool. The model included fixed effects for activity type (domestic, occupational, recreational, trade/visit), and was adjusted for age class, sex, and location and duration.

Posterior densities represent marginal distributions of estimated effects, with the vertical dashed line indicating the null (log = 0).

**Table S6.** Self-reported reasons for not taking part in the last mass drug administration.

**Table S7.** Self-reported participation in the last MDA by age group.

**Table S8.** Frequency of Lake Victoria travel and distribution of reported activities, stratified by duration in the lake.

