## Supplementary Data for "Local habitual movement as a mechanism for *Schistosoma mansoni* transmission resurgence – a causal analysis"

### Supplementary Material

#### *Village selection*

To select villages for the main study a baseline survey was carried out in twenty villages which were all approximately five kilometres from Lake Victoria, Uganda. This distance was chosen as previous evidence suggested that villages over five kilometres from Lake Victoria were at significantly lower risk of infection with prevalence below 15% (1). For baseline surveillance only the WHO guidelines for rapid assessment were followed; approximately fifty school-aged children (SAC) were sampled in each village (2). Each child provided a single stool sample, which was examined using one Kato Katz thick smear slide (3). Based on these initial surveys, five villages were selected for the main study which had all reached the WHO target of elimination of transmission which resulted in communities who had either low

(<10%) or moderate (<50%) prevalence. Simulated power analyses were conducted during this process to ascertain the required sample sizes in each population https://github.com/iamjessclark/travelStudySchisto

In each village prior to study commencement a meeting was hosted by the Ministry of Health team to which all members of the community were invited. During this meeting the purpose and design of the study was explained and any questions answered.

**Table S1.** Participants responses to what activity they did at Lake Victoria put into groups for statistical analysis.

| **Activity response** | **Grouping** |
|---|---|
| Fetch water for home use | Domestic |
| Fetch water for work use | |
| Water | |
| Washing | |
| Wash | |
| Going to the garden | |
| Cultivating | |
| Cross bypassing | |
| Boarding a boat | |
| Go to work | Occupation |
| Fishing | |
| Ferrying bricks | |
| Swimming | Recreation |
| Recreation | |
| Play | |
| Motorcycle | |
| Bathing | |
| Visit relative | Trade/Visit |
| Visit friends | |
| Trade | |
| Sell jackfruits | |
| To buy food | |
| Selling ripes | |
| Sell cassava | |
| School | |

| |
|---|
| Prayers |
| Operating small drinking place |
| Market |
| Friends |
| For vegetables |
| Food |
| Fishmonger |
| Buying fish |
| Buying clothes |
| Buy silverfish |

Model validation and restructuring

The structure of the DAG was validated against the observed data by testing the implied conditional independencies between the variables. The implied independencies were as follows: age was independent of sex; duration independent of MDA if conditioned on age, location and travel or activity, age, location and sex; MDA independent from sex when conditioned on age, location and travel. To identify and test these implied independencies, we use the concept of d-separation where two variables are d-separated given a set of conditioning variables if all paths between them are "blocked". A path is blocked if it contains a collider (a node with two arrows pointing towards it, e.g. A → B ← C) that is not conditioned on (nor any of its descendants), or if it contains a fork (e.g. A ← B → C) that is conditioned on (Pearl et al., 2016). This testing was carried out by constructing a general linear model (GLM) in R, using the appropriate link function, with the outcome as the response variable and the exposure the predictor (corresponding to the direction of the assumed causal influence in the DAG). Additional variables were included in the model based on the conditioning rules outlined above. Independence between the variable and the outcome was assumed if the p-value between the outcome and the exposure variable from the GLM was >0.1.

If a conditional independence did not hold in the data, the DAG structure was updated to include additional plausible paths or modify existing ones. This process was repeated until the DAG was both biologically meaningful and all implied conditional independencies were consistent with the observed data. This final validated DAG was then used to identify appropriate adjustment sets for estimating both total and direct effects across the causal framework.

The final structure of the DAG was as follows: Age and sex were exogenous variables with no parent nodes, therefore influencing downstream behavioural and exposure related variables. Age and sex class were treated as temporally prior to location based on the assumption that demographic characteristics influence where individuals live (4). Location was exogenous to all other variables to capture spatial heterogeneity in exposure risk. The relationship between travel frequency and infection had both indirect and direct effects. This relationship was indirectly mediated by activity type as those who travel more frequently are more likely to engage in regular activities like chores (domestic) or work (occupation) (5), and also mediated by duration, this was decided by causal priority (4), since individuals must first choose to travel to the lake before they enter it. A direct path from travel frequency to infection was also included to capture potential exposure mechanisms not fully explained by duration or activity, such as unmeasured behavioural factors. Activity type influenced duration, reflecting that certain activities involve longer exposure (5–7). Duration in the lake was a direct exposure variable that causally influenced infection (8,9). MDA history was assumed to influence infection, as treatment reduces worm burden (10). Travel frequency was also assumed to have a negative effect on MDA as those who travel often may not be present at their home village when the MDA took place (11).

**Table S2.** Minimal adjustment sets used in total and direct effect models by exposure.

| | **Minimal adjustment set** | |
|---|---|---|
| **Exposure** | **Total effect** | **Direct effect** |
| Travel frequency | Age, Sex, Location | Age, Sex, Location, MDA and Activity |
| Activity | Age, Sex, Location and Travel frequency | Age, Sex, Location, Travel frequency and Duration |
| MDA | Age, Location and Travel frequency | Age, Location and Travel frequency |
| Duration | Age, Sex, Location and Activity | Age, Sex, Location and Activity |

<u>Sensitivity analysis of priors for the coefficients</u>

For all Bayesian analyses, we used weakly informative priors, as there was little to no prior data available to support more specific assumptions. To assess the sensitivity of our findings to prior choice, we also fitted models with narrower coefficient priors: Normal(0, 5) and Normal(0, 2) (Table S3). The most notable difference under these more informative priors was a reduction in the estimated effect of activity on egg burden (EPG) (Figure S1). However, the relative ranking and direction of effects across activity categories remained consistent, with domestic activity showing the strongest effect. This effect remained strongly supported, as its posterior distribution did not cross the null (Figure S1A). Under the more restrictive (0, 2) priors, although the pattern of relative effects remained unchanged, the increased regularisation introduced by the tighter prior led to greater uncertainty around the estimated effects, and all posterior distributions crossed the null, including that of domestic activity (Figure S1B).

This is likely due to the relatively small number of individuals with positive egg counts, which limits the information available to overcome prior shrinkage. Despite this, we believe that the use of weakly informative priors remains justified, given the exploratory nature of the analysis and the absence of prior knowledge in this setting.

Using narrower coefficient priors made no meaningful difference when estimating the effect of travel frequency, MDA nor duration in the lake.

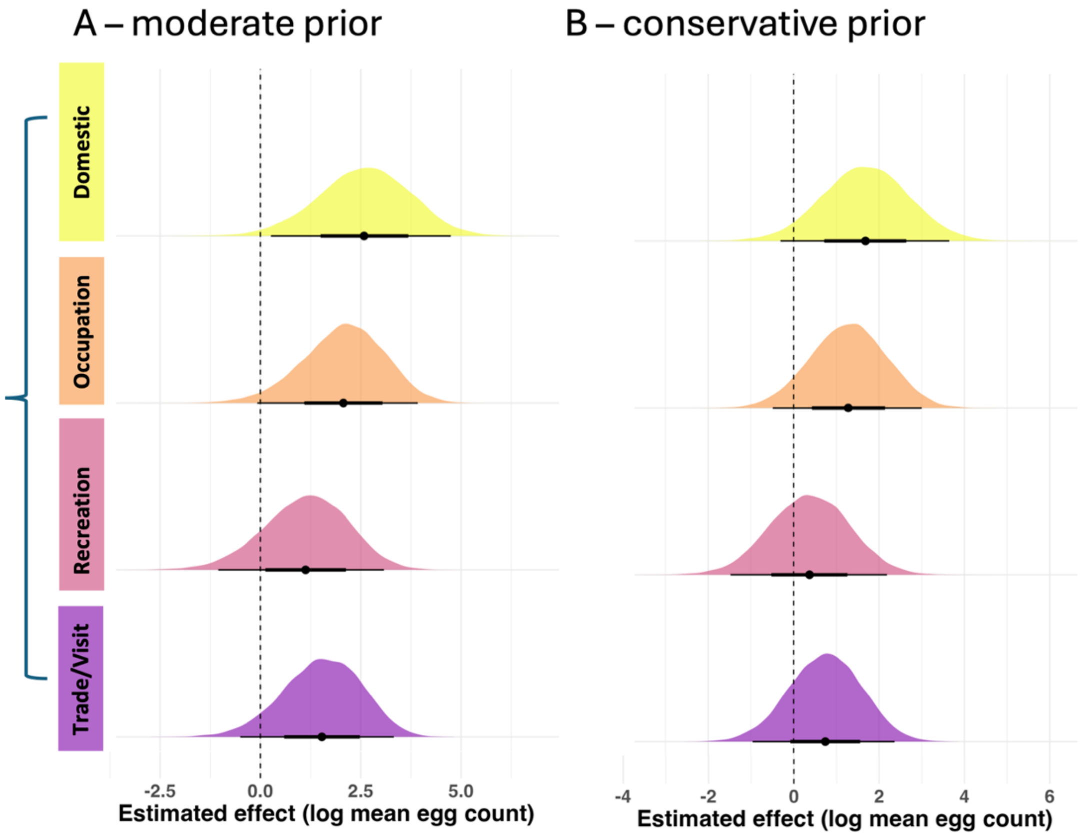

**Figure S1.** Total effects of activity on *Schistosoma mansoni* infection burden.
Posterior estimated total effects of lake-related activities, on the log mean of *S. mansoni* eggs per gram of stool. The model included fixed effects for activity type (domestic, occupational, recreational, trade/visit), and was adjusted for age class, sex, and location and duration.
Posterior densities represent marginal distributions of estimated effects, with the vertical dashed line indicating the null (log = 0).

A) Moderately informative coefficient priors of Normal(0,5)
B) Conservative coefficient priors of Normal(0,2)

**Table S3.** Model parameters and prior distributions.

| Model | Parameter | Prior | Notes |
|---|---|---|---|
| Bernoulli (Infection status) | Intercept | Normal(1.5, 2.5) | Expected baseline infection probability of ~18% |
| Bernoulli (Infection status) | Regression coefficients | Normal(0, 10) | Weakly informative, no strong prior knowledge |
| Gamma (EPG) | | Normal(2.5, 1) [log scale] | Approximates log mean of observed egg counts |
| Gamma (EPG) | Regression coefficients | Normal(0, 10) | Weakly informative, allows potentially large effects |
| Gamma (EPG) | Shape parameter | Exponential(1) | Allows for overdispersion |
| Sensitivity Analysis | Regression coefficients - moderate | Normal(0, 5) | Moderate prior, intercept and exponential as above |
| Sensitivity Analysis | Regression coefficients - conservative | Normal(0, 2) | Conservative prior, intercept and exponential as above |

**Table S4.** *Schistosoma mansoni* prevalence and mean eggs per gram of stool (EPG) ordered by travel frequency to Lake Victoria, Uganda

| Travel frequency | Age class | Number of participants | Infected• (%) | Mean EPG* (standard deviation) |
|---|---|---|---|---|
| None | Adult | 115 | 28% | 8.6 (10.5) |
| None | SAC | 65 | 28% | 45.0 () |
| None | PSAC | 76 | 36% | 12.8 (11.5) |
| Just once | Adult | 24 | 29% | 51.0 (81.9) |
| Just once | SAC | 18 | 33% | 4.4 (3.7) |
| Just once | PSAC | 8 | 25% | 0.5 () |
| Less than once per month | Adult | 1 | 0% | 0 |
| Less than once per month | SAC | 2 | 50% | 0.2 () |
| Once per month | Adult | 29 | 45% | 5.7 () |
| Once per month | SAC | 10 | 70% | 6.6 (2.2) |
| Once per month | PSAC | 10 | 40% | 5.0 () |
| Twice per month | Adult | 10 | 60% | 9.4 (5.1) |
| Twice per month | PSAC | 1 | 100% | 0.2 () |
| Once per week | Adult | 15 | 40% | 15.8 (11.1) |
| Once per week | SAC | 24 | 33% | 8.3 (10.0) |
| Once per week | PSAC | 17 | 24% | 13.7 () |
| Twice per week | Adult | 11 | 55% | 5.1 (5.5) |
| Twice per week | SAC | 10 | 50% | 4.4 (4.0) |
| Twice per week | PSAC | 1 | 100% | 0 |
| Daily | Adult | 61 | 44% | 12.5 (14.4) |
| Daily | SAC | 60 | 40% | 9.4 (13.9) |
| Daily | PSAC | 17 | 35% | 0 |

•using a positive Kato-Katz or a G-score ≥3, *excluding zeros and only using Kato-Katz

**Table S5.** Frequency of Lake Victoria travel and distribution of reported activities, stratified by age group, sex, and village.

| | | | | | Activity (percentage of all those that reported travel to the Lake in this population) | | | | | | | |
|---|---|---|---|---|---|---|---|---|---|---|---|---|
| **Characteristic** | **n** | **Total days travelled in last 3 months (Percentage of population)** | | **Average number of days** | **Domestic** | | **Occupation** | | **Recreation** | | **Trade/Visit** | |
| Adult | 266 | 150 | (56) | 42 | 57 | (38) | 21 | (14) | 7 | (5) | 59 | (40) |
| SAC | 189 | 122 | (65) | 50 | 42 | (34) | 7 | (6) | 32 | (26) | 35 | (29) |
| PSAC | 130 | 54 | (42) | 35 | 26 | (48) | 0 | (0) | 9 | (17) | 13 | (24) |
| Female | 364 | 203 | (56) | 43 | 85 | (42) | 9 | (4) | 20 | (10) | 80 | (40) |
| Male | 220 | 123 | (56) | 45 | 40 | (33) | 19 | (15) | 28 | (23) | 27 | (21) |
| B | 119 | 75 | (63) | 35 | 37 | (49) | 8 | (11) | 24 | (32) | 6 | (8) |
| Nw | 133 | 77 | (58) | 46 | 8 | (10) | 8 | (10) | 18 | (23) | 29 | (38) |
| Nam | 111 | 79 | (71) | 68 | 71 | (90) | 1 | (1) | 1 | (1) | 6 | (8) |
| Nan | 91 | 59 | (65) | 23 | 6 | (10) | 8 | (14) | 4 | (7) | 39 | (66) |
| S | 131 | 39 | (56) | 35 | 3 | (8) | 3 | (8) | 1 | (3) | 27 | (69) |
| **Total** | **585** | **326** | **(56)** | **43** | **125** | **(38)** | **28** | **(9)** | **48** | **(15)** | **107** | **(33)** |

*PSAC: preschool age children, SAC: school age children*

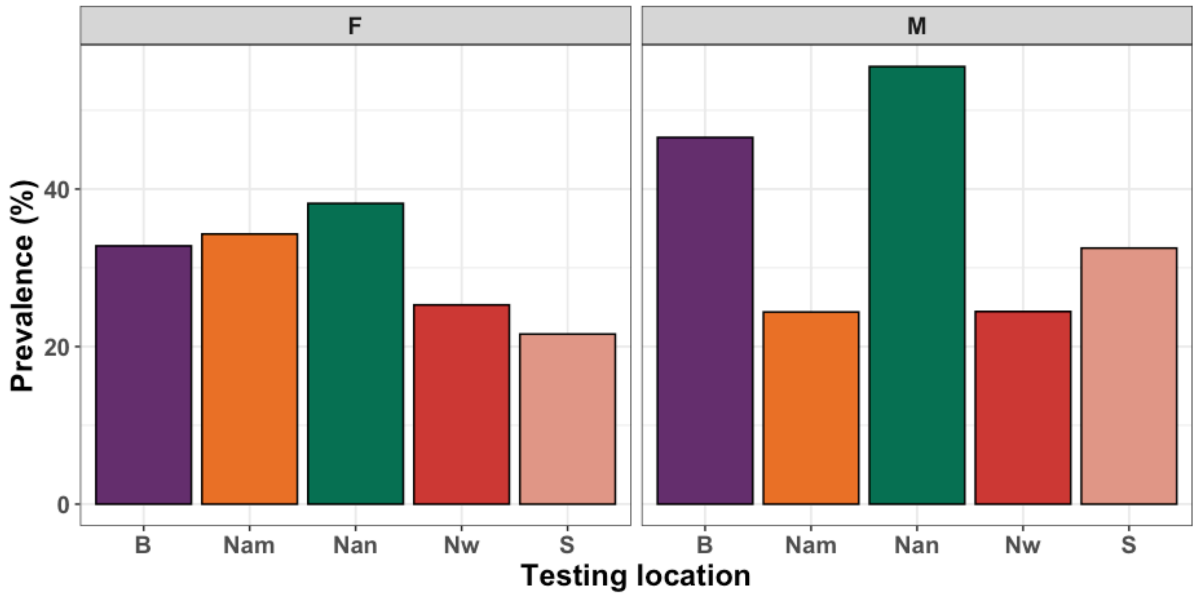

**Figure S2.** Prevalence of *S. mansoni* by village and Sex. Prevalence measured by a positive Kato Katz from three duplicate slides or if a POC-CCA test had a G-score ≥3. F = female and M = male.

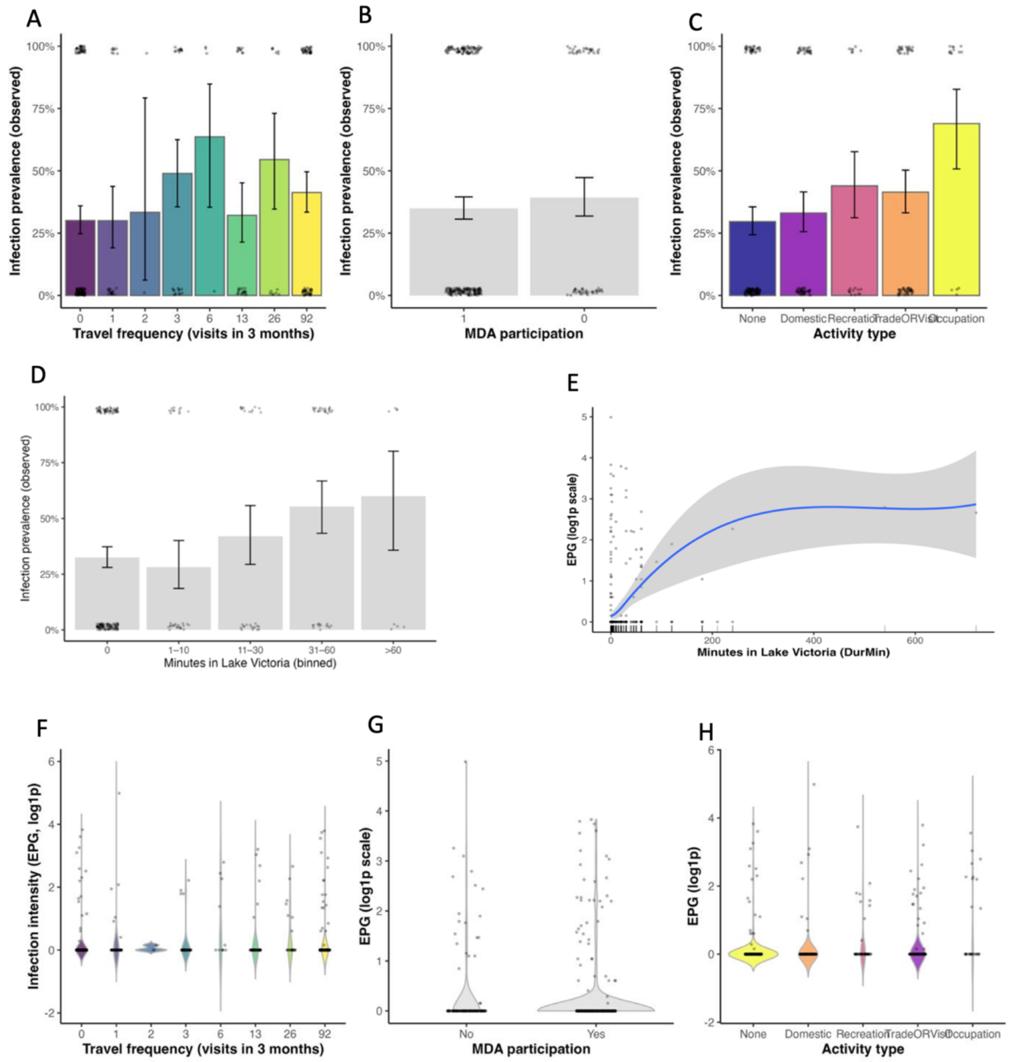

**Figure S3. Observed infection prevalence and intensity by behavioural and exposure variables.**

Observed infection prevalence (top and middle rows) and infection intensity (bottom row) plotted against behavioural and exposure variables included in the structural causal model. Bars and ribbons show observed mean prevalence with 95% binomial confidence intervals. Points represent individual infections (1 = infected, 0 = uninfected), shown with jitter for visibility.

**(A)** Infection prevalence by travel frequency to Lake Victoria (visits in the past three months).
**(B)** Infection prevalence by participation in the most recent mass drug administration (MDA).
**(C)** Infection prevalence by activity type at Lake Victoria.
**(D)** Infection prevalence by time spent in the lake (binned durations).
**(E)** Infection intensity (EPG, $\log_{1+}$ scale) by time spent in the lake (continuous).

**(F–H)** Infection intensity (EPG, $\log_{1+}$ scale) by travel frequency (F), MDA participation (G), and activity type (H).

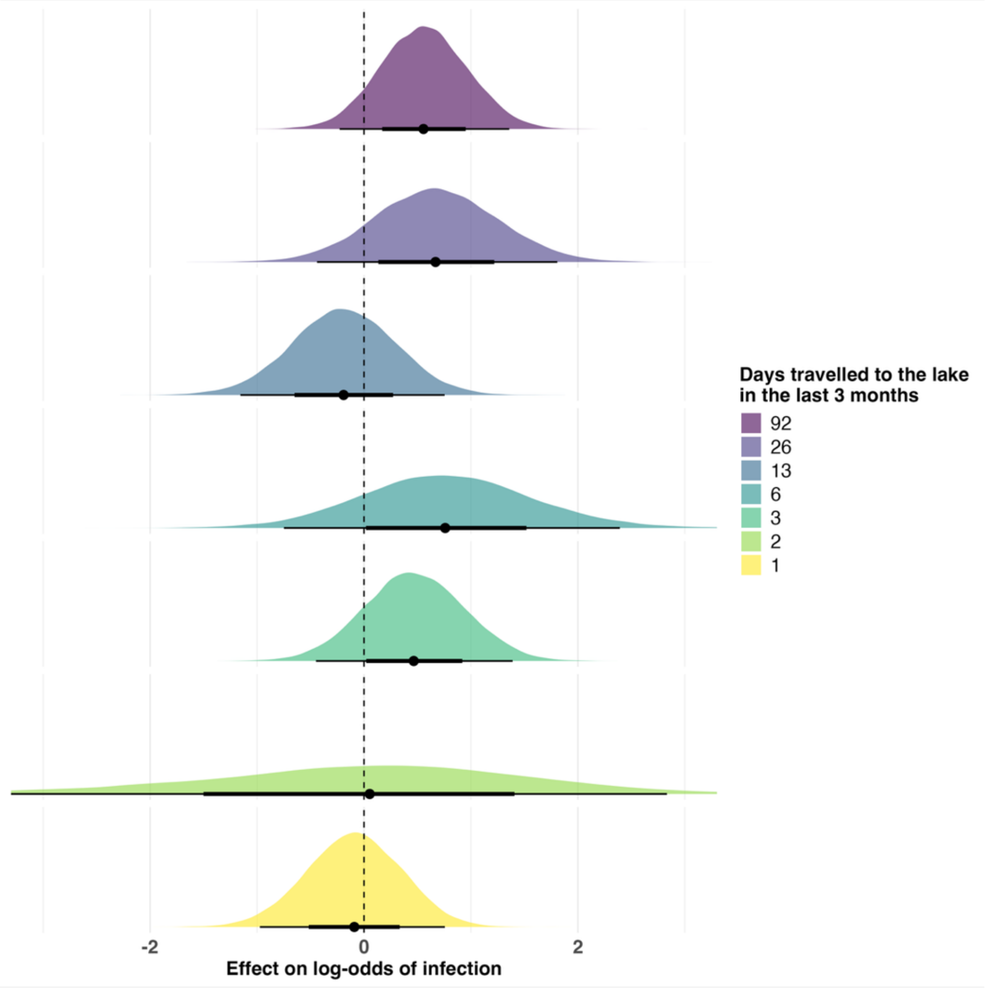

**Figure S4.** Direct effect of travel frequency on log odds of infection with *Schistosoma mansoni.*

Posterior distributions for the estimated direct effects of self-reported travel frequency to Lake Victoria on the probability of infection, relative to individuals reporting no travel in the preceding three months (reference category). Estimates are log-odds coefficients from a Bayesian logistic regression model with a Bernoulli likelihood and logit link function. The model included fixed effects for travel frequency, age class, sex (binary), and location and the mediator's activity and MDA. Densities represent marginal posterior distributions for each travel frequency level, with the vertical dashed line indicating the null effect (log-odds = 0). Travel frequency levels are ordered from highest (92 days) to lowest (1 day) reported visits to the lake.

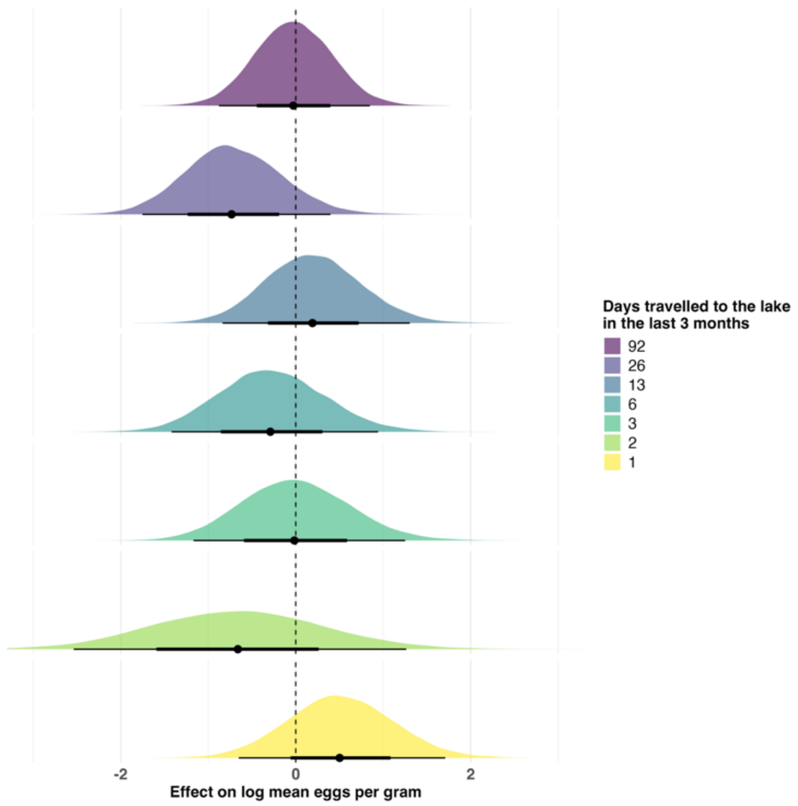

**Figure S5.** Direct effect of travel frequency on log mean on burden of *Schistosoma mansoni* eggs per gram in stool.

Posterior distributions for the estimated direct effects of self-reported travel frequency to Lake Victoria on mean eggs per gram of stool, relative to individuals reporting no travel in the preceding three months (reference category). Estimates are log mean coefficients from a Bayesian logistic regression model with a gamma likelihood and log link function. Densities represent marginal posterior distributions for each travel frequency level, with the vertical dashed line indicating the null effect (log-odds = 0). Travel frequency levels are ordered from highest (92 days) to lowest (1 day) reported visits to the lake.

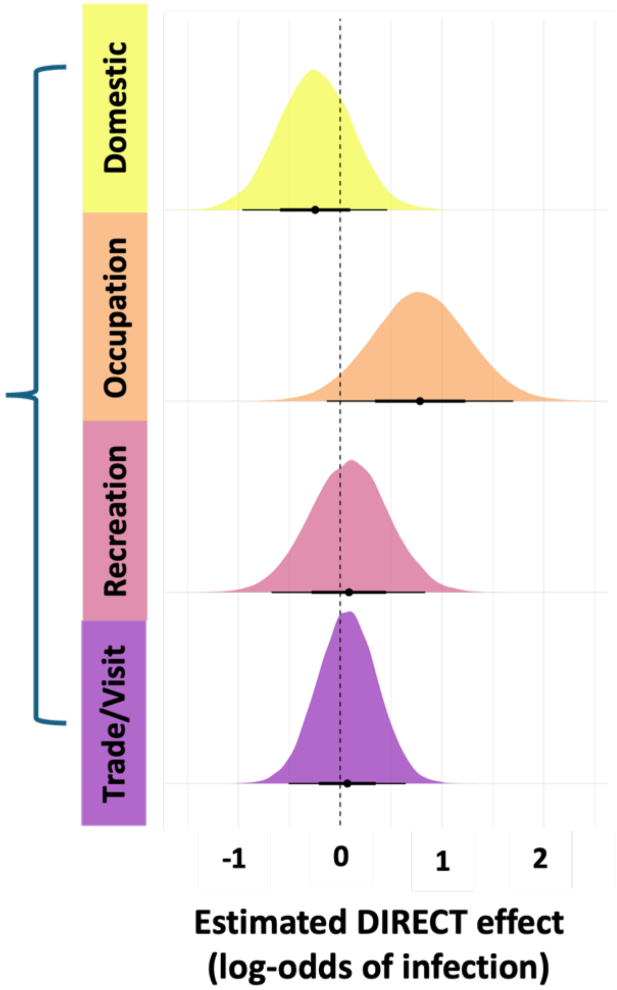

**Figure S6.** Direct effects of activity on *Schistosoma mansoni* infection probability. Posterior distributions for the estimated direct effects of lake-related activities, the log odds of infection. The model included fixed effects for activity type (domestic, occupational, recreational, trade/visit), and was adjusted for age class, sex, and location and duration. Posterior densities represent marginal distributions of estimated effects, with the vertical dashed line indicating the null (log = 0).

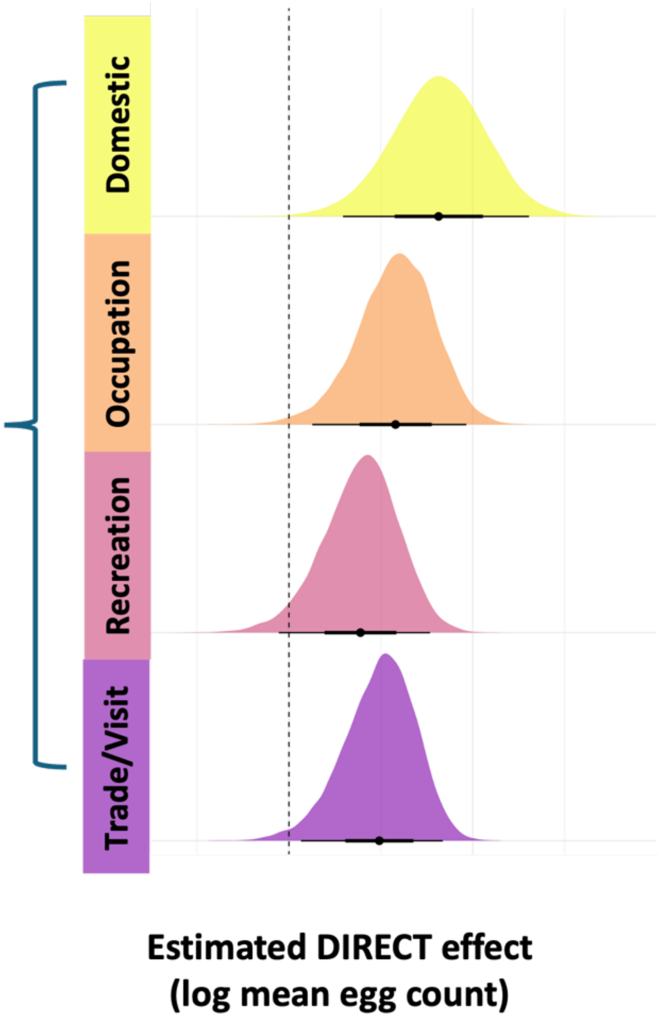

**Figure S7.** Direct effects of activity on *Schistosoma mansoni* infection burden.
Posterior distributions for the estimated direct effects of lake-related activities, on the log mean of *S. mansoni* eggs per gram of stool. The model included fixed effects for activity type (domestic, occupational, recreational, trade/visit), and was adjusted for age class, sex, and location and duration.
Posterior densities represent marginal distributions of estimated effects, with the vertical dashed line indicating the null (log = 0).

**Table S6.** Self-reported reasons for not taking part in the last mass drug administration.

| Reason for not taking part | Number of people |
|---|---|
| Too young | 42 |
| Away | 28 |
| Did not know about it | 22 |
| Not offered it | 15 |
| Did not trust it / concerned about side effects | 13 |
| COVID affected decision | 8 |
| Not infected, so did not need it | 3 |
| Did not know why | 19 |

**Table S7.** Self-reported participation in the last MDA by age group.

| Age group | Total population size | % who reported taking part in last MDA |
|---|---|---|
| PSAC | 130 | 50% |
| SAC | 189 | 82% |
| Adults | 266 | 77% |

**Table S8.** Frequency of Lake Victoria travel and distribution of reported activities, stratified by duration in the lake.

| Travel frequency | Activity | n | Proportion of those in travel frequency group | Average duration in the lake – minutes (sd) |
|---|---|---|---|---|
| Never | None | 232 | 91% | 0 () |
| | Domestic | 5 | 2% | 0 (0) |
| | Occupation | 1 | 0% | 2 () |
| | Recreation | 2 | 1% | 40 (28) |
| | TradeORVisit | 16 | 6% | 7 (22) |
| Just once | None | 12 | 24% | 1 (3) |
| | Domestic | 6 | 12% | 6 (7) |
| | Occupation | 4 | 8% | 24 (27) |

| | | | | |
|---|---|---|---|---|
| | Recreation | 12 | 24% | 25 (25) |
| | TradeORVisit | 16 | 32% | 18 (53) |
| Less than once a month | Domestic | 1 | 33% | 0 () |
| | TradeORVisit | 2 | 67% | 30 (0) |
| Once a month | None | 1 | 2% | 20 () |
| | Domestic | 13 | 27% | 8 (18) |
| | Occupation | 6 | 12% | 45 (23) |
| | Recreation | 2 | 4% | 40 (28) |
| | TradeORVisit | 27 | 55% | 20 (34) |
| Twice a month | Occupation | 4 | 36% | 215 (237) |
| | TradeORVisit | 7 | 64% | 1 (4) |
| Once a week | None | 1 | 2% | 5 () |
| | Domestic | 28 | 50% | 39 (49) |
| | Occupation | 2 | 4% | 38 (11) |
| | Recreation | 9 | 16% | 40 (57) |
| | TradeORVisit | 16 | 29% | 40 (37) |
| Twice a week | None | 2 | 9% | 1 (1) |
| | Domestic | 4 | 18% | 45 (90) |
| | Occupation | 3 | 14% | 260 (399) |
| | Recreation | 5 | 23% | 52 (18) |
| | TradeORVisit | 8 | 36% | 22 (28) |
| Daily | None | 5 | 4% | 5 (8) |
| | Domestic | 73 | 53% | 5 (12) |
| | Occupation | 9 | 7% | 64 (70) |
| | Recreation | 20 | 14% | 29 (23) |
| | TradeORVisit | 31 | 22% | 17 (24) |

11. Adriko M, Faust CL, Carruthers LV, Moses A, Tukahebwa EM, Lamberton PHL. Low praziquantel treatment coverage for Schistosoma mansoni in Mayuge district, Uganda,

due to the absence of treatment opportunities, rather than systematic non-compliance. Trop Med Infect Dis. 2018;3(4):111.
